# BRAVE-NET: Fully Automated Arterial Brain Vessel Segmentation In Patients with Cerebrovascular Disease

**DOI:** 10.1101/2020.04.08.20057570

**Authors:** Adam Hilbert, Vince I. Madai, Ela M. Akay, Orhun U. Aydin, Jonas Behland, Jan Sobesky, Ivana Galinovic, Ahmed A. Khalil, Abdel A. Taha, Jens Wuerfel, Petr Dusek, Thoralf Niendorf, Jochen B. Fiebach, Dietmar Frey, Michelle Livne

## Abstract

**Introduction:** Arterial brain vessel assessment is crucial for the diagnostic process in patients with cerebrovascular disease. Noninvasive neuroimaging techniques such as time-of-flight (TOF) magnetic resonance angiography (MRA) imaging are applied in the clinical routine to depict arteries. They are, however, only visually assessed. Fully automated vessel segmentation integrated into the clinical routine could facilitate the time-critical diagnosis of vessel abnormalities and might facilitate the identification of valuable biomarkers for cerebrovascular events. In the present work, we developed and validated a new deep learning model for vessel segmentation, coined BRAVE-NET, on a large aggregated dataset of patients with cerebrovascular diseases.

**Methods:** BRAVE-NET is a multiscale 3-D convolutional neural network (CNN) model developed on a dataset of 264 patients from 3 different studies enrolling patients with cerebrovascular diseases. A context path, dually capturing high- and low-resolution volumes, and deep supervision were implemented. The BRAVE-NET model was compared to a baseline Unet model and variants with only context paths and deep supervision, respectively. The models were developed and validated using high-quality manual labels as ground truth. Next to precision and recall, the performance was assessed quantitatively by Dice coefficient (DSC); average Hausdorff distance (AVD); 95- percentile Hausdorff distance (95HD) and via visual qualitative rating.

**Results:** The BRAVE-NET performance surpassed the other models for arterial brain vessel segmentation with a DSC = 0.931, AVD = 0.165 and 95HD = 29.153. The BRAVE-NET model was also the most resistant towards false labelings as revealed by the visual analysis. The performance improvement is primarily attributed to the integration of the multiscaling context path into the 3-D Unet and to a lesser extent to the deep supervision architectural component.

**Discussion:** We present a new state-of-the-art of arterial brain vessel segmentation tailored to cerebrovascular pathology. We provide an extensive experimental validation of the model using a large aggregated dataset encompassing a large variability of cerebrovascular disease. The framework provides the technological foundation for improving the clinical workflow and can serve as a biomarker extraction tool in cerebrovascular diseases.

## Introduction

Stroke is one of the leading causes of death worldwide and 15 million people each year suffer a stroke (WHO EMRO | Stroke, Cerebrovascular accident | Health topics). Stroke is a cerebrovascular disease and as such is characterized by changes in the arterial vasculature of the brain such as stenosis and occlusion of vessels. Information about the arterial vessel status proves crucial for the diagnostic process. In chronic cerebrovascular disease - which is often present prior to stroke - it can serve as a biomarker: It was shown that brain vessel status can predict the likelihood of further stroke events (Gutierrez Jose et al., 2015). In the acute clinical setting, brain vessel status provides stroke physicians with pivotal information. For example, acute arterial vessel occlusions qualify patients for mechanical thrombectomy, the best reperfusion therapy of stroke at the moment (Turc et al., 2019). This explains the high clinical relevance to depict brain vessels in cerebrovascular disease.

Noninvasive neuroimaging techniques are used to depict brain vessels. One method is magnetic resonance (MR) time-of-flight (TOF) imaging. It is fast, has no ionizing radiation exposure and can depict the arterial vasculature in high detail. Clinical reading of vessel imaging is based on visual judgment alone, as there is a lack of fully automated vessel segmentation methods. Fully automated segmentation means in the clinical context that the segmentation results would be rapidly available on the scanner console without the need of extra post-processing. Extensive, time-consuming and non-standardized image postprocessing is an important obstacle to widespread clinical application. Fully automated analysis of TOF images could facilitate the diagnosis of vessel abnormalities and allow the quantification of the cerebrovascular status, e.g. arterial vessel density or arterial vessel diameters, to make potential biomarkers for diagnostic stratification easily available (Dengler et al., 2016; Santos et al., 2016; Yoo et al., 2018; Dutra et al., 2019; Murray et al., 2019).

Convolutional neural networks (CNNs) - a type of artificial neural networks (ANN) tailored for image analysis - have become the method of choice for vessel segmentation including the brain vasculature and showed promising results in recent applications (Moccia et al., 2018; Tetteh et al., 2018; Livne et al., 2019a). However, these previous studies suffer from various impediments that still limit their usability in the clinical setting. First, post-processing of the images such as noise filtration and brain masking are commonly applied (Passat et al., 2005; Livne et al., 2019a). The necessity of image processing may not only lead to suboptimal generalization due to a lack of standardized processing methods but may disqualify the models for clinical use due to time constraints, as the same processing needs to be applied to each image before predictions can be run. Second, the studies are often performed on small datasets or even simulated data due to a lack of large amounts of high-quality labeled data. Third, a lack of data heterogeneity limits the generalizability of these models to new datasets. Lastly, the suggested frameworks are rarely developed in cases with vessel pathologies. These constraints constitute a severe challenge for the clinical application of the models, since it may impair their ability to accurately detect vessels and vessel abnormalities in patients with cerebrovascular pathologies.

Recognizing this challenge, this work presents a high performance fully automated framework for vessel segmentation in patients with cerebrovascular disease addressing all the above-mentioned limitations, coined BRAVE-NET (BRAinVEssel-NETwork). BRAVE-NET is a multiscale 3-D CNN model designed to dually capture high- and low-resolution volumes thus enabling enhanced distinction of brain vessels from other structures such as skull areas and arteries not feeding the brain, making masking and other post-processing steps obsolete. The model was developed and validated using high-quality labeled data derived from multiple datasets of patients (n=264) with cerebrovascular disease. This approach ensured the high performance and high generalization of BRAVE-NET.

## Methods

### Data

#### Accessibility

Due to data protection laws, the imaging data used in this study cannot be published at the current time point. Implementation of the proposed network, as well as the training, prediction and evaluation framework can be found on Github at https://github.com/prediction2020/brain-vessel-segmentation.

#### Patients

Retrospective data from the PEGASUS (Martin et al., 2015), 7UP (Ultrahigh-Field MRI in Human Ischemic Stroke – a 7 Tesla Study) and 1000Plus (Hotter et al., 2009) studies, with 264 patients in total, were analyzed in this study.

From the PEGASUS and 7UP studies, 74 and 9 patients with chronic steno-occlusive disease were available for this study, respectively. From the 1000Plus study, we included 181 patients with acute stroke. All mentioned studies were carried out in accordance with the recommendations of the authorized institutional ethical review board of Charité-Universitätsmedizin Berlin (PEGASUS and 1000Plus) and the Berlin state ethics boards (7UP). All subjects gave written informed consent in accordance with the Declaration of Helsinki.

#### Imaging properties

Time-of-Flight (TOF) Magnetic Resonance Angiography (MRA) images were used to train all benchmarked models. TOF relies on the fact that, within an imaged volume, inflowing blood has high magnetization compared to stationary tissue that becomes magnetically saturated by multiple radiofrequency pulses. TOF-MRA is one of the most important methods for non-contrast neurovascular and peripheral MRA.

In the PEGASUS and 1000Plus studies, TOF imaging was performed on a Magnetom Trio 3T whole-body system (Siemens Healthcare, Erlangen, Germany) using a 12-channel receive radiofrequency (RF) coil (Siemens Healthcare) tailored for head imaging. In the 7UP study, TOF imaging was performed on a Magnetom Verio 3T whole body system (also Siemens Healthcare) using a 12-channel receive radiofrequency (RF) coil (Siemens Healthcare) tailored for head imaging.

Parameters of the TOF imaging of each dataset are shown in Table 1.

**Table 1.** Time of flight (TOF) magnetic resonance imaging (MRI) parameters.

| Dataset | Voxel size | Matrix size | TR/TE (ms) | Time of acquisition (min:sec) | Flip angle (degrees) |
| --- | --- | --- | --- | --- | --- |
| PEGASUS | 0.52 x 0.52 x 0.65 | 312 x 384 x 127 | 22 / 3.86 | 3:50 | 18 |
| 7UP | 0.28 x 0.28 x 0.6 | 644 x 768 x 136 | 24/ 3.60 | 5:54 | 18 |
| 1000Plus | 0.52 x 0.52 x 0.65 | 312 x 384 x 127 | 22 / 3.86 | 3:50 | 18 |

#### Data labeling

Intra-cerebral arteries as well as the major brain-supplying arteries, i.e. the internal carotid arteries, the vertebral arteries and the basilar artery were labeled. For PEGASUS and 7UP data, ground-truth labels of the arterial brain vessels were generated semi-manually using a standardized pipeline. Pre-labeling of the vessels was performed by a thresholded region-growing algorithm using the *regiongrowingmacro* module implemented in MeVisLab (Mevis Medical Solutions, Bremen, Germany). To tackle inter-rater variability in label generation, these pre-labeled data were thoroughly manually corrected by either OUA and EA (both junior raters) and then cross-checked by the other rater. Junior raters were trained for image segmentation and were only allowed to independently label images once their performance met ground truth standards. These labels were then checked subsequently by VIM (9 years experience in stroke imaging). Thus, each ground-truth was eventually checked by 3 independent raters, including 1 senior rater. The total labeling time with this framework amounted to 60–80 min per patient. For 1000Plus data, the MeVisLab-based pre-segmentation step was replaced by using a 2D Unet segmentation model, developed in earlier work (Livne et al., 2019a). These pre-segmentations were - as described above - manually corrected either by OUA, EA or JB, cross-checked by another junior rater and then subsequently checked by VIM. For this approach, the total labeling time was reduced to 40-60 minutes per patient.

#### Data pre-/post-processing

No pre- or post-processing methods were applied in the presented framework that are necessary to be performed on datasets prior to any prediction. Importantly, no mask was used for validation or test. All the developed models can operate on raw TOF images coming directly from the scanner and are not dependent on any processing step or external tool.

For the training step only, and here to reduce the training time of the models, a heuristical mask was automatically computed for each TOF image to exclude training patches containing only air as follows: 1) The images were smoothed with a 16×16×16 averaging filter in a shifting window fashion 2) A threshold of intensity values at 10 was applied (intensity > 10 implies ROI). This provides an automatically generated mask for the training phase which contains all the brain tissue but also a thick border area around the skull. The inclusion of patches from these border areas proved to be crucial for the learning process, particularly for the distinction of the skull and the ability to capture the distinctive neighborhood of small vessels in that area.

#### Dataset preparation and patch extraction

Images from each dataset were used for training, model selection and evaluation. A 4-fold cross-validation methodology was employed to ensure the robustness of all models towards different training and test sets. Therefore data was split randomly into 4 distinct training and test sets making sure that each set contained the same number of patients from each source dataset (Pegasus, 7UP, 1000Plus). Furthermore, 15% of images were separated randomly without overlaps from each training set for validation and used for parameter selection. Figure 1. illustrates the creation of training, validation and test sets. Final number of patients was 170, 29, 65 in each set respectively; including equal numbers of patients from each source across folds.

**Figure 1.**
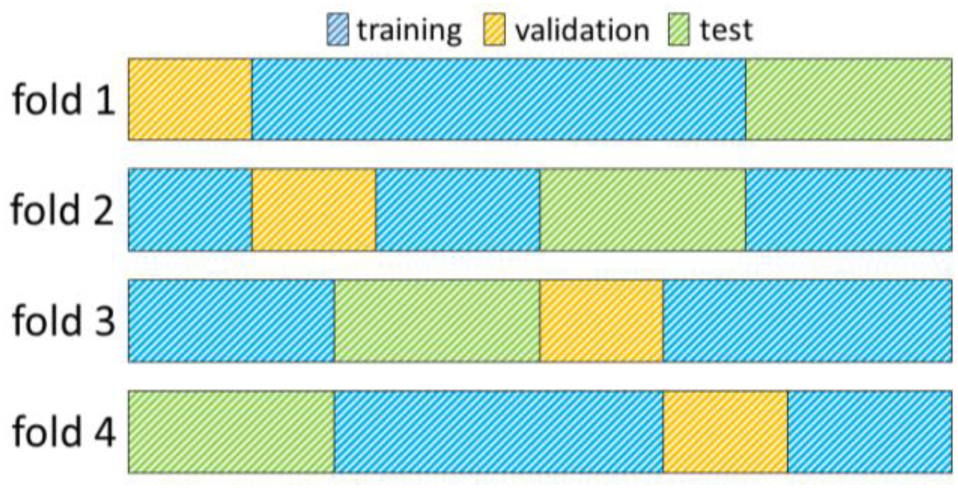
Illustration of a 4-fold cross-validation framework. Each row in the figure represents a fold. Data assigned to the training-validation- and test sets are indicated with the colors blue, yellow and green respectively. In each fold, the training set comprises 60%, the validation set 15% and the test set 25% of all images (170, 29, 65 images, respectively).

The task of brain vessel segmentation on 3D MRI images challenges deep learning techniques from several aspects. First, processing of whole brain volumes at once requires significant resources in terms of GPU memory. Second, the distribution of vessels compared to brain tissue is very sparse. The physiological arterial vascular volume fraction of the brain is 1.5%. TOF-detectable arterial vessel voxels can be as low as on average 0.3% of all voxels within the brain. Finally, sufficient training of deep neural networks requires numerous images, while our dataset is - like most medical imaging datasets - limited in size. To solve these challenges, we redesigned the segmentation task as voxel-wise classification and trained our models on patches that represent an arbitrary neighborhood of the center voxel. Accordingly, 2000 locations per TOF image (i.e. patient) were randomly sampled within the heuristical mask, ensuring 50% of the samples were vessel-centric. Patches of sizes [64×64×8] and [128×128×16] were extracted around each location, and comprised the input patches of our proposed framework.

### Segmentation frameworks

#### Baseline Unet

The backbone of the proposed framework is realized by an adjusted Unet architecture (Ronneberger et al., 2015) with 4 levels, shown on Figure 2. The Unet model consists of an encoding path (left side), and a decoding path (right side). At each level of the encoding, we employ a sequence of Convolution, Rectified Linear Unit (ReLU) activation and Batch-normalization two times consecutively followed by Max-pooling (Ioffe and Szegedy, 2015). Additional dropout is applied in the first sequence after the Batch-normalization (Srivastava et al., 2014). We set the kernels and stride to 3×3×3 and 1 for convolutions and 2×2×2 and 2 for max-pooling. The resolution and depth of feature maps are reduced to half and doubled respectively after each level. As an extension of the original Unet architecture, we add 2 consecutive fully connected layers to the last level realized by 1×1×1 convolutions.

**Figure 2.**
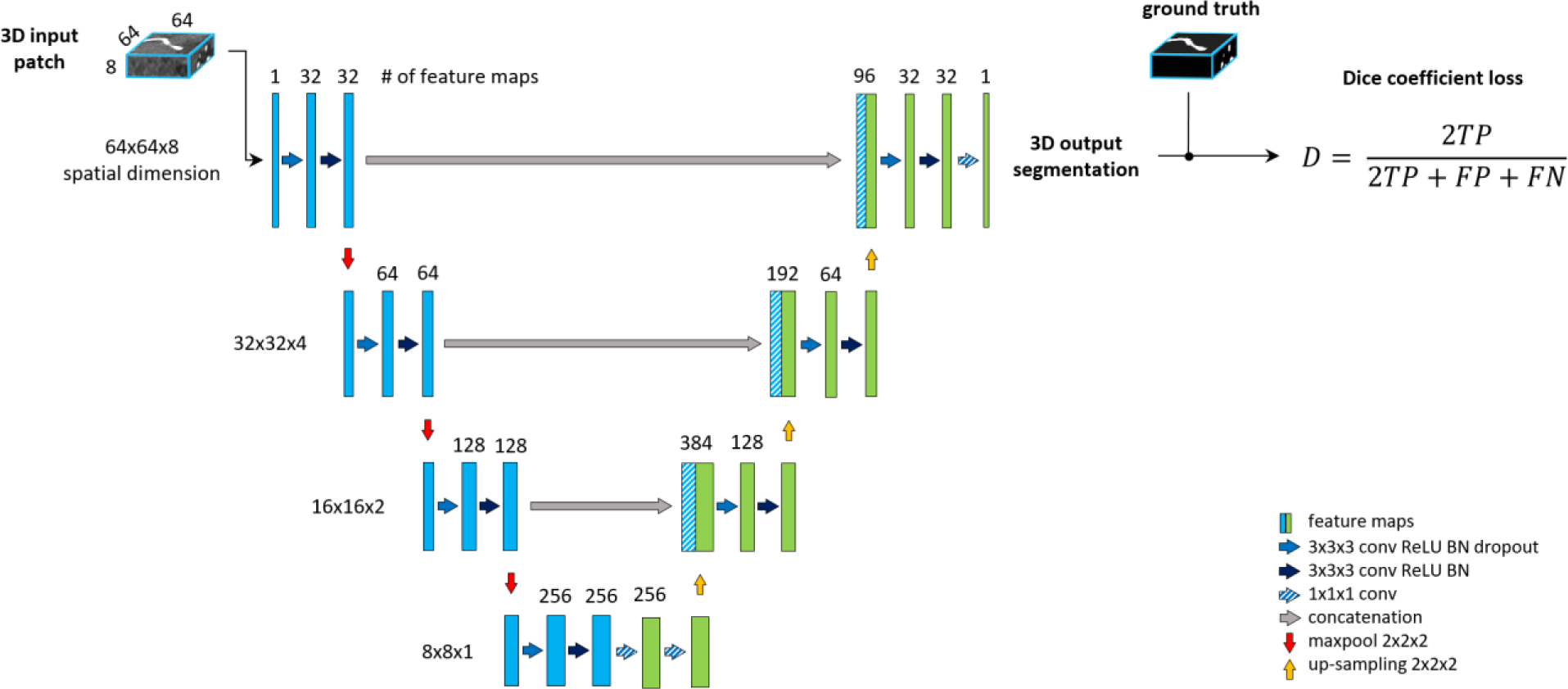
The baseline architecture. A standard Unet architecture, coined *Unet*, consisting of 4 levels, 2 consecutive sequences of convolution, ReLU, Batch-normalization at each level of encoding (left side) and decoding (right side) part. 2 additional fully connected layers are added to the last level realized by 1×1×1 convolutions.

The decoding part of the architecture recovers the original input dimensions by applying the same sequences of Convolution, ReLU, Batch-normalization but replacing max-pooling with up-sampling at each level. Additionally, corresponding feature maps from the encoding part are concatenated to the input of each decoding level. Finally, 1×1×1 convolution with sigmoid activation is applied to map feature maps into a binary prediction map, which is used to calculate DSC loss.

#### Context Unet

Patch-wise formulation of the segmentation problem comes at the cost of neglecting the full spatial context of input images. In theory, the model should learn to distinguish vessels from other structures within the brain. On the other hand, as described under the data labeling section, we are only interested in segmenting specific arterial brain vessels which have specific spatial locations. Furthermore, Livne et al. showed the limitation of a standard Unet variant in detecting small vessels (Livne et al., 2019a). A significant amount of these small vessels lie close to the skull i.e. has a particularly distinctive spatial context. To tackle these problems, a multiscale approach is applied (Choi et al.; Kamnitsas et al., 2017; Yue et al., 2019). We extend the base architecture with a context aggregation under the hypothesis that it will improve the discrimination between vessels of interest and other constructs and particularly improve the segmentation of small vessels.

This approach is depicted in Figure 3. The encoding part of our Unet is extended with a so-called context path. The input of this path is a larger - context - patch, extracted around the same center voxel as for the other encoding path. Inspired by Kamnitsas et al. 2017 (Kamnitsas et al., 2017), the input patch is then down-sampled by average-pooling with 2×2×2 kernels and stride of 2, i.e. to the same dimension but half-resolution compared to the other encoding path. The down-sampling allows for neglecting fine details and focusing on contextual information. The down-sampled input is fed into a parallel, equivalent sequence of layers. The two parallel downward paths are realized as duplicates (i.e. with no shared parameters) in order to enable distinctive feature encodings for the context and original patch. The output of the encoding paths - i.e. bottom level - are concatenated and fed through 2 fully connected layers realized by 1×1×1 convolution followed by ReLU. Finally, the residuals of each level of both encoding paths are concatenated to the input of the corresponding decoding level to facilitate the contribution of spatial and context information in the final prediction map.

**Figure 3.**
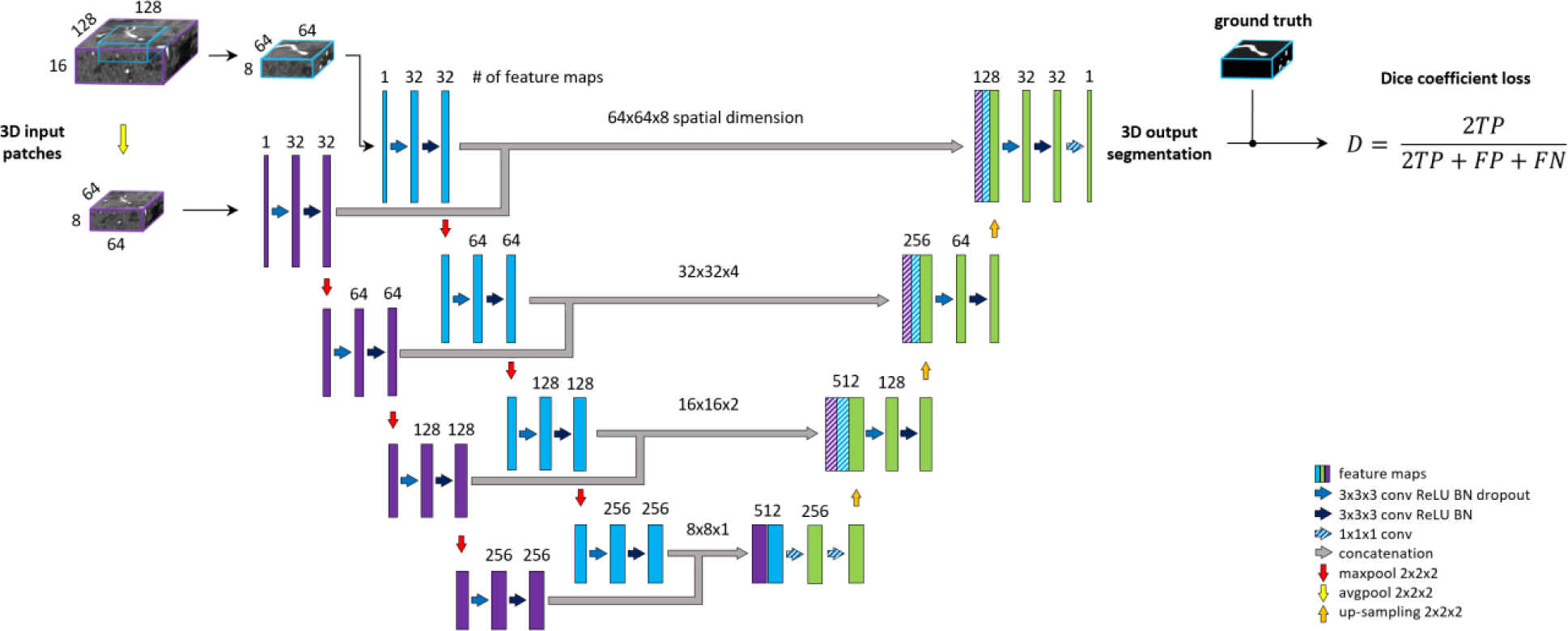
Unet architecture extended with context aggregation, coined *Context-Unet*. Additional context path (purple columns) is added to the encoding (left side) part. The initial downsampling of the 128×128×16 input patch is followed by the same number of layers as the other encoding path (blue columns). The two paths are concatenated and fed into the decoding part (right side). Input patches are extracted around the same center voxel.

#### Deep Supervision Unet

Deep supervision is a method commonly used to avoid the problem of exploding or vanishing gradients in deep networks by forcing intermediate layers to produce more discriminative features. Stawiaski (Stawiaski, 2017) implemented deep supervision by down-sampling ground truth segmentations and weighting each coefficient equally in the loss function. Other implementations aggregated the coarse, low-resolution feature maps into the final convolutional layers and thus incorporated different features in a single loss coefficient (Chen et al., 2014; Long et al., 2014; Folle et al., 2019). In contrast, we aim to facilitate the convergence of intermediate layers by direct supervision. Feature maps from intermediate decoding levels (i.e. all except bottom and final level) are first up-sampled to the output dimension and then fed into a 1×1×1 convolutional layer with sigmoid activation to produce prediction masks. From each output of the model, DSC loss is computed with respect to the ground truth labeling. The loss coefficients are weighted and summed to create the training loss of the framework. We aim to reflect more emphasis on the final prediction thus assign 0.5 weight to the final layer and distribute the remaining 0.5 across intermediate outputs. A deep supervision variant of the architecture is coined here as DS-Unet. An illustration of deep supervision integration in the final architecture can be found in Figure 4.

**Figure 4.**
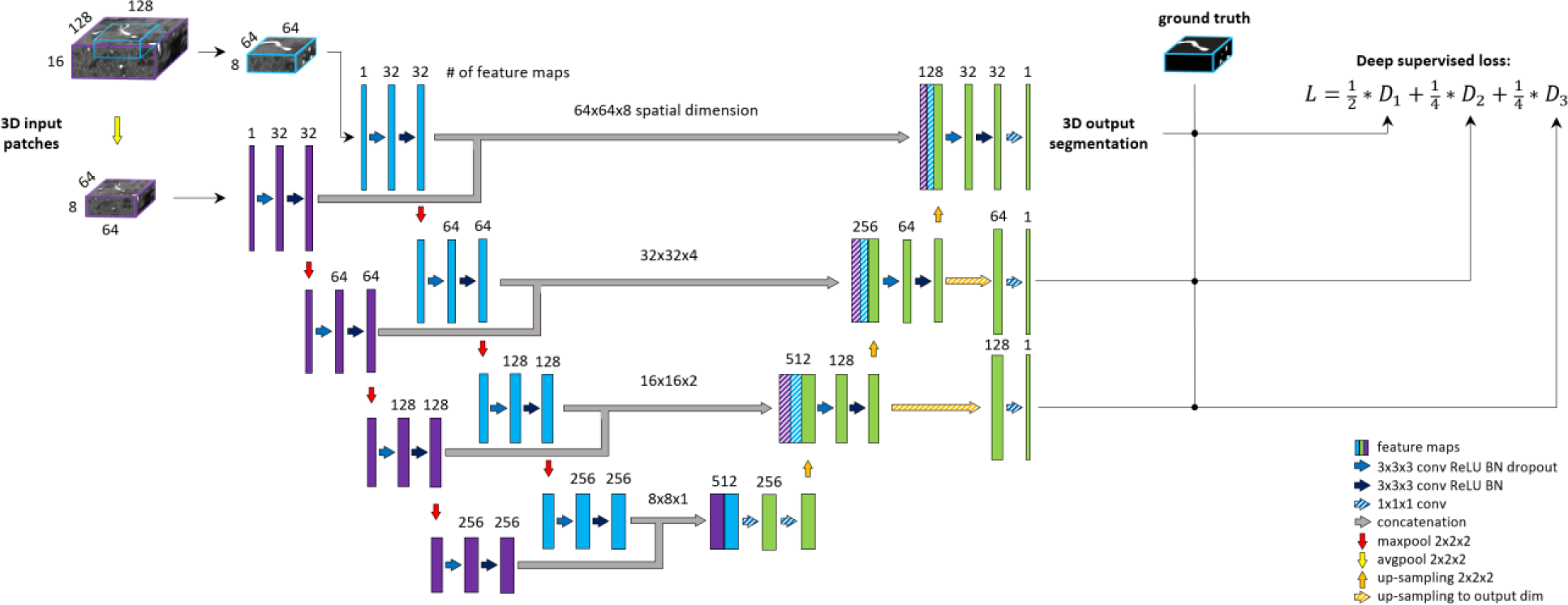
BRAVE-NET architecture. The model is composed of a Unet architecture extended with context aggregation and deep supervision. Feature maps of intermediate decoding levels are up-sampled to output - spatial - dimension (striped yellow arrows) and prediction masks are produced by 1×1×1 convolution and sigmoid activation (striped blue arrows).

#### BRAVE-NET

The proposed BRAVE-NET architecture integrates the 3D Unet with both approaches introduced above, multiscaling and deep supervision. Figure 4 shows the BRAVE-NET architecture - including an illustration of the integration of deep supervision to the loss function. A model card outlining the model characteristics of BRAVE-NET can be found in the appendix, as suggested by Mitchell et al. (Mitchell et al., 2019).

The number of trainable parameters are shown in Table 2. Naturally, Context-Unet and BRAVE-NET contain more parameters due to the multi-scale encoder part, while DS-Unet has only few more trainable parameters than Unet due to the 2 additional final convolutions of the up-sampled outputs.

**Table 2.** Number of trainable parameters of all implemented models.

|  | Unet | DS-Unet | Context-Unet | BRAVE-NET |
| --- | --- | --- | --- | --- |
| number of trainable parameters | ~6 million | ~6 million | ~10 million | ~10 million |

### Model training and evaluation

#### Experimental setup

The models were trained in the above-described cross-validation framework. The optimal learning rate of the optimization process was selected by highest average performance across validation sets evaluated on whole brain segmentation. The largest batch-size feasible was used to fully utilize computational resources and to optimize training time. Models were trained on a high performance deep learning server using a single NVIDIA Titan RTX GPU.

To analyze the contribution of the proposed extensions, different configurations of the segmentation framework were tested. The performance of BRAVE-NET was compared to the DS- Unet, the Context-Unet and to the baseline Unet.

#### Training scheme

The designated input (BRAVE-NET: [64×64×8], [128×128×16], DS-Unet: [64×64×8], Context- Unet: [64×64×8], [128×128×16], Unet: [64×64×8]) and ground truth patches ([64×64×8]) were used to train the models. All models were implemented in Python using Tensorflow Deep learning framework (Abadi et al.) and were trained using Adam optimizer (Kingma and Ba, 2014).

The final layer of each model utilizes a voxel-wise sigmoid activation function, defined as *p*(*x*) =*x*/(*x* + *exp*(*f*(*x*))), where *f*(*x*) denotes the output value of the last [1×1×1] convolutional layer at the voxel position *x ∈ Ω* with *Ω ∈ Z*^3^and *p*(*x*) is the approximated probability of a voxel *x* to be a vessel. The same activation is applied to each output of the deep supervision method. DSC loss was used to update the model parameters and to monitor training convergence, throughout all experiments. DSC between two binary volumes - i.e. prediction and ground truth - is defined as *D* =2*TP*/(2*TP* + *FP* + *FN*), where *TP* is the number of true positive voxels, *FP* is the number of false-positive voxels and *FN* is the number of false-negative voxels. Further derived:

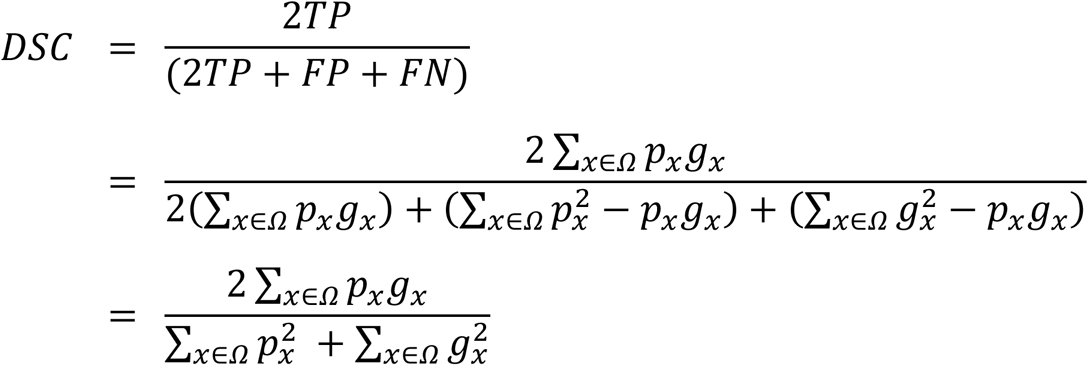

Where *p*_*x*_ *∈ P*: *Ω* → {0,*x*} is the binary prediction volume and *g*_*x*_ *∈ G*: *Ω* → {0,*x*} the binary ground truth volume. This yields in the following differentiated gradient at each position *j*:

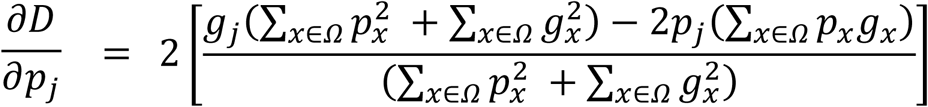

We monitored - patch-level - validation loss during training and employed EarlyStopping with a patience of 3 epochs to optimize training time and to save the best model with respect to validation loss. The weight of every convolutional layer was initialized with the commonly used Glorot uniform initialization scheme (Glorot and Bengio, 2010).

#### Performance evaluation

The performance of vessel segmentation models can be assessed from many different angles. As (Moccia et al., 2018) also highlights, different anatomical regions or imaging modalities might require different measures to be considered. To give a broader view of the performance of our models, we report five different metrics: Precision, Recall, DSC, 95 percentile Hausdorff Distance (95HD) and Average Hausdorff Distance (AVD). We included Precision and Recall to compare models by the quality and completeness of segmentations respectively and DSC, 95HD, and AVD to assess the spatial overlap between predictions and ground truth. The evaluation metrics were calculated on whole-brain segmentations (reconstructed from patch-wise predictions), by using the open-source evaluation tool from (Taha and Hanbury, 2015). We report mean values and standard deviation across test sets defined by the cross-validation framework.

#### Visual assessment

For qualitative assessment, the predicted segmentation masks were transformed by in-house python code where true positives (TP), false positives (FP), and false negatives (FN) were assigned different voxel values (True negatives (TN) remained labeled with 0). The images were then visualized by overlaying these new masks with the original scans using ITK-Snap software (Yushkevich et al., 2006). By adjusting the opaqueness, it was possible to qualitatively assess which structures were correctly identified and which anatomical structures dominated with errors. Owing to the time burden of reviewing such a large number of images (4 models, 264 patients, 4 folds), only one fold was visually assessed. The first fold was chosen without prior inspection of any results. The 65 patients in the fold were assessed based on a predefined scheme slightly adapted from Livne et al (Livne et al., 2019a). Large vessels were defined as all parts of the internal carotid artery (ICA), the basilar artery, the anterior cerebral artery (ACA) and the M1, and P1 segments of the middle and posterior cerebral artery. All other parts were considered small vessels. The results of the visual analysis are summarized qualitatively in the results section. The scheme was the following: Vessel pathology detected (yes/no); Large vessels, overall impression (bad, sufficient, good); Small vessels, overall impression (bad, sufficient, good); Large vessels, errors (FP or FN dominating; or balanced); Small vessels, errors (FP or FN dominating, or balanced); Other tissue type segmentation errors (yes/no).

## Results

The BRAVE-NET model yielded the highest performance on all measures except Recall, namely DSC of 0.931, AVD of 0.165, 95HD of 29.153 and Precision of 0.941. Highest Recall of 0.930 was achieved by the Context-Unet model. The standard Unet baseline achieved a DSC of 0.928, AVD of 0.232, 95HD of 33.259, Precision and Recall of 0.928 and 0.929 respectively. The detailed test results of the cross-validation framework are presented in Table 3. Example segmentation results can be found in Figure 5.

**Table 3.** Test results of cross-validation for all models, average (SD) over all folds are given. Best overall performance was achieved by our proposed BRAVE-NET model.

| Models (all 3D) | DSC (SD) | AVD (SD) | 95HD (SD) | Precision (SD) | Recall (SD) |
| --- | --- | --- | --- | --- | --- |
| Unet | 0.928 (0.004) | 0.232 (0.041) | 33.259 (1.060) | 0.928 (0.005) | 0.929 (0.004) |
| DS-Unet | 0.927 (0.004) | 0.222 (0.033) | 33.420 (0.866) | 0.927 (0.003) | 0.929 (0.005) |
| Context-Unet | 0.931 (0.004) | 0.198 (0.067) | 29.279 (1.900) | 0.934 (0.006) | **0.930 (0.008)** |
| BRAVE-NET | **0.931 (0.003)** | **0.165 (0.013)** | **29.153 (0.988)** | **0.941 (0.005)** | 0.923 (0.004) |

**Figure 5.**
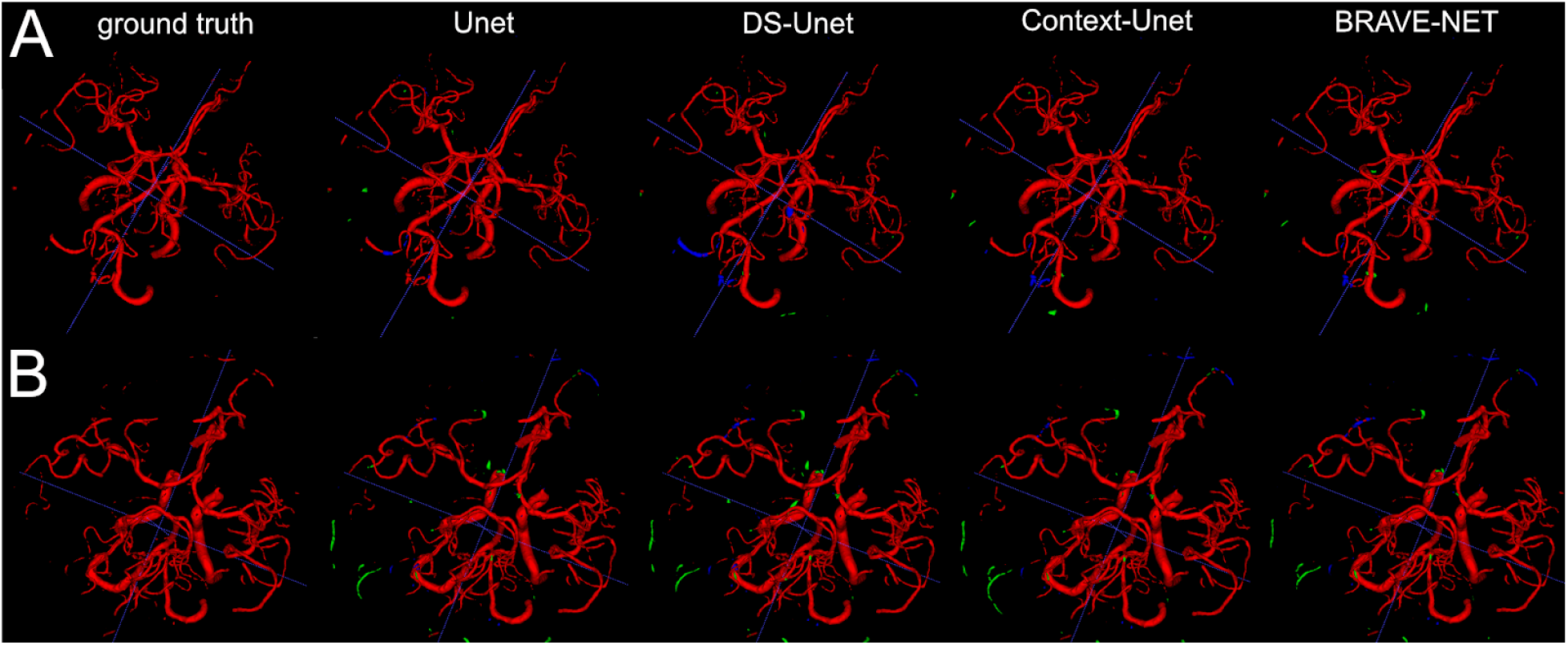
As demonstrated by the high performance results, segmentation results were excellent. Two example patients are shown in rows A and B, showing in the first column the ground truth and in the following the segmentation results derived from the Unet, the DS-Unet, the Context-Unet and finally the BRAVE-NET model. Only very few false positive (green) and false negative (blue) vessels can be observed.

Context-aggregation alone was shown to improve the baseline performance of Unet for each measure while deep supervision alone only improved the AVD measure. Combining both techniques however yielded the best overall performance.

For all models, the visual analysis found comparable excellent performance. Of the 64 patients, 15 showed visible steno-occlusive changes. These vessel pathologies were detected by all models. The overwhelming majority of images was rated to have good performance both for large vessels (Unet: 95%; DS-Unet: 93%; Context-Unet: 95%; BRAVE-NET: 93%) as well as small vessels (Unet: 91%; DS-Unet: 91%; Context-Unet: 92%; BRAVE-NET: 92%). Large vessel error was dominated by false positives (approx. 65% of images for all models), whereas small vessel error was dominated by balanced error (approx. 63% of images for all models).

Besides excellent vessel segmentation performance, the visual inspection revealed a high rate of falsely labeled rare pathologies and other tissue classes. The false-labeling-rate was lowest for the BRAVE-NET model (40%), followed by Context-Unet (43%), DS-Unet (49%) and Unet (50%) with the highest false-labeling-rate. Figure 6 demonstrated the improved performance of the BRAVE-NET model. Of all found false labelings, the majority were partially labeled segments of the external carotid artery in the neck region (48%), followed by false positively labeled deep veins (29%), as illustrated in Figure 7. Other false labelings were rarer (false labeling of hyperintensities in the mesencephalon 10%; cortical laminar necrosis 6%, cerebellar structures 2%, arteria meningea media 2%, proteinaceous fluid (2%) and false negative labeling of rete mirabile structures 2%).

**Figure 6.**
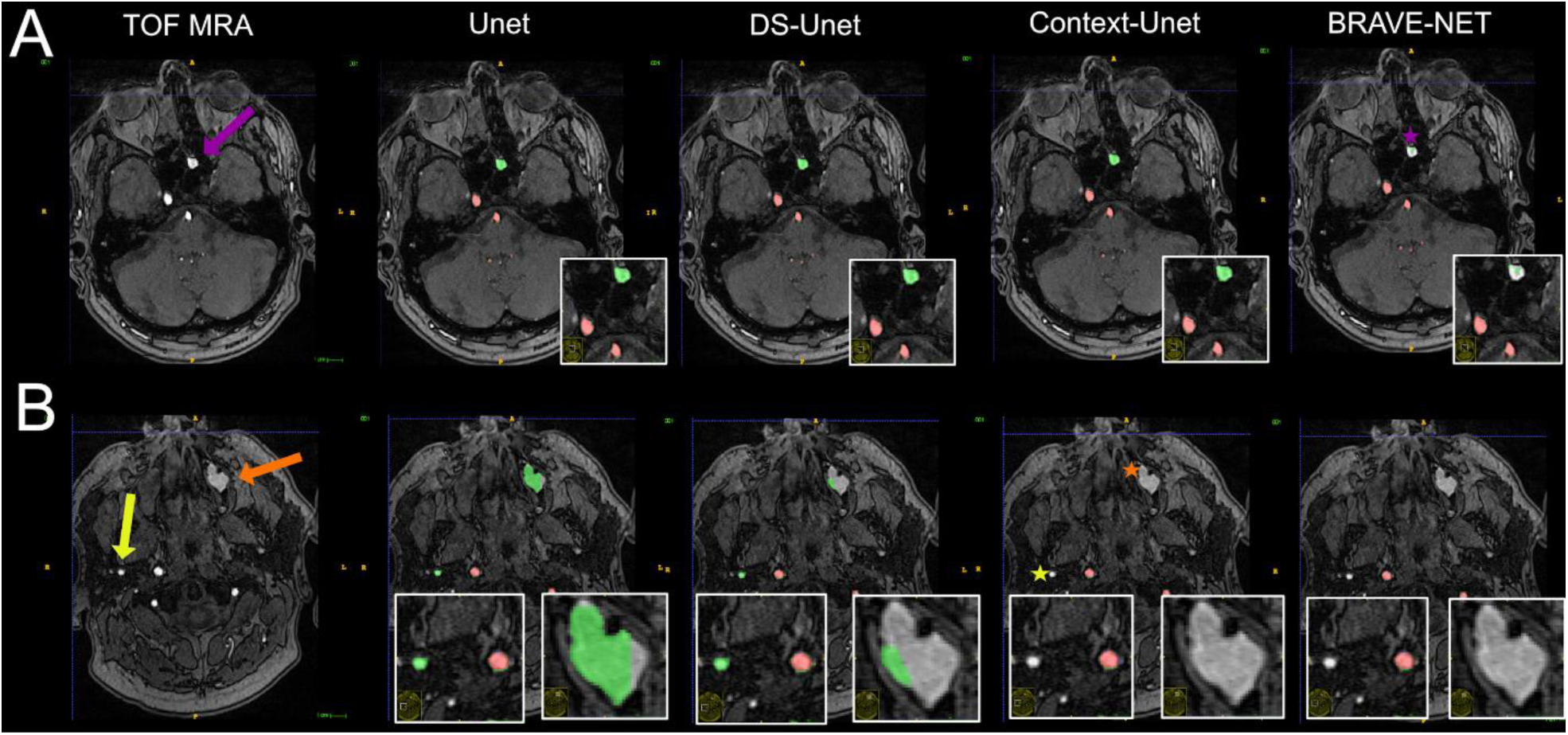
Vessel segmentation of an example patient. Two slices are shown (A and B). True positive and false positive are shown in red and green respectively. The patient had hyperintense proteinaceous fluid in the sinuses (purple and orange arrows) indicative of chronic sinusitis and an extranial vessel (yellow arrow), which were all false positively labeled in the plain Unet (column 2). It is evident that the network needs high contextual anatomic knowledge to distinguish the hyperintense roundish sinusitis structures from vessels. Also, this example illustrates how close - anatomically - extracranial vessels can be located to brain-supplying vessels (correctly labeled by all networks). For the structure labeled by the purple arrow, only the BRAVE-NET network correctly classifies the majority of voxels as non-vessels (purple star). For the larger structure which is less hyperintense and further away from brain areas as well as the extracranial vessel, the Context-net alone correctly classified the structures (orange and yellow asterisks). This example illustrates that while the overall differences between the models do not seem large, in certain pathological cases the contextual information proves crucial to avoid false positive segmentation results.

**Figure 7.**
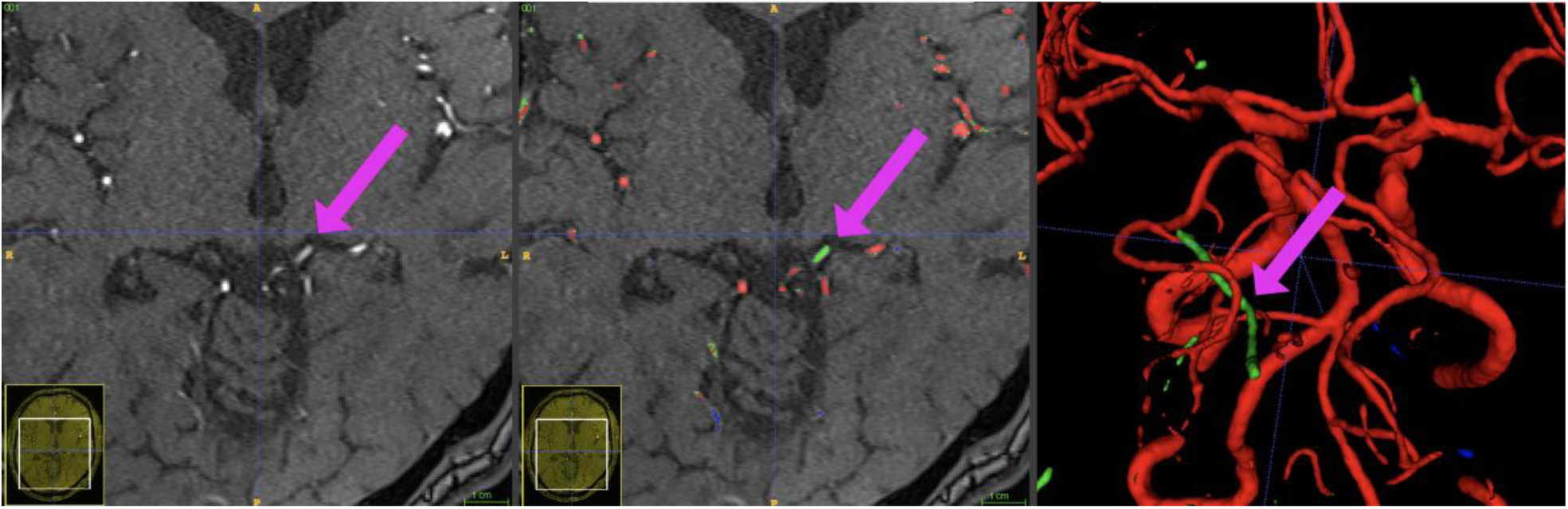
Exemple patient showing the phenomenon of a hyperintense Vena basalis rosenthali. In some patients, the flow of venous structures feeding the sinus rectus is high enough to be depicted in TOF-imaging. Since these structures are located very centrally in the brain and are very close to arterial structures, none of the presented models was able to correctly ignore these structures in all images. Blue crosshairs depict the same coordinates in all three images.

## Discussion

We present the BRAVE-NET model for fully automated arterial vessel segmentation from TOF-MRA images in cerebrovascular disease. The model combines multiscaling and deep-supervision as extensions of the standard Unet approach and yielded a new state-of-the-art performance. This was achieved by robust validation using the largest presented dataset in the literature so far. BRAVE-NET is suitable for clinical application, as it can be applied on raw images without the necessity for additional image processing. The superiority of the BRAVE-NET was demonstrated quantitatively and was confirmed via visual assessment. It outperformed the other tested models especially in cases of visible pathologies and false labeling. BRAVE-NET marks a significant step towards increased applicability of deep learning based segmentations in the clinical setting and the continued development of quantitative vascular biomarkers in cerebrovascular disease.

Reported models in the literature for brain vessel segmentation are usually limited to a DSC of 0.90 (Babin et al., 2013; Wang et al., 2015b; Chen et al., 2017; Livne et al., 2019a). Notable exceptions, besides our work, are the works of Ni et al. and Tatel et al. who reported DSC values of 0.96 and 0.94, respectively (Ni et al., 2020; Patel et al., 2020). They were, however, obtained for different modalities, CT angiography and digital subtraction angiography, respectively. Moreover, the DSC measure alone is not sufficient to provide the full picture regarding the needs of clinical application. Here, our model has other major advantages that have to our knowledge not been achieved together in one single model so far.

First, the majority of MRA-based vessel segmentation frameworks presented so far in the literature require image pre-processing such as downsampling, brain masking, intensity correction, image normalization, and various other methods before they can be applied (Lesage et al., 2009; Chen et al., 2017; Livne et al., 2019a; Ni et al., 2020; Taher et al., 2020). Applications in computed tomography angiography (CTA) do not differ in that regard and include for example deformable matching, atlas co-registration, candidate vessel selection and feature extraction (Passat et al., 2005; Meijs et al., 2017). In contrast, our framework completely avoids time- and resource-consuming pre- and post-processing methods that rely on domain knowledge. Moreover, this means that the BRAVE-NET model is usable on the scanner console in real-time given that a full brain prediction for clinically used dimensions - such as in the 1kplus study - can be available within minutes on a standard CPU system (AMD Ryzen 7 1700X), in our case 2 minutes. As comparison, predictions for the same images on a standard GPU (NVIDIA Titan Xp) system take 40 seconds.

Second, previous frameworks were developed using smaller datasets ranging from 10 to 100 patient scans derived from a single scanner (Passat et al., 2005; Wang et al., 2015b; Chen et al., 2017, 3; Livne et al., 2019a; Ni et al., 2020; Patel et al., 2020). Taher et al. used a patient cohort size (N=270) similar to our study (Taher et al., 2020).

Third, the majority of reported models are based on data obtained from healthy individuals and did not include patients with (vessel) pathologies. Exceptions are the work of Meijs et al. who developed CTA-based models on data of patients with a suspected stroke (Meijs et al., 2017), the work of Patel et al. including patients with aneurysms (Patel et al., 2020) and the work of Livne et al. who developed MRA-based models on patients with cerebrovascular disease and additionally tested the generalization of the model on datasets from different scanners (Livne et al., 2019a). The present work extends the latter work by adding a large number of patients from an additional data source allowing development, validation and testing on a combined multi-scanner, high quality labeled dataset. Furthermore, the model was developed on data with a broad range of pathologies present in patients with acute and chronic cerebrovascular disease. Thus, not only a broad variety of occlusions and stenoses were present, but also important pathologies such as cortical laminar necrosis, rete mirabile, and others. Such a property is of high importance in the field of deep learning, as many applications suffer from bias due to constrained and homogenous data selection (Ho et al., 2019; Hofmanninger et al., 2020). Consequently, such models usually perform worse on datasets differing from the dataset distribution used for training. For example, Taher et al. used the freely available model of Livne et al. for vessel segmentation (Livne et al., 2019a, 2019b) on their own data and noticed a reduced performance compared to the original publication (Taher et al., 2020). Here, the increased generalizability of the model presented in this work is supported by its ability to capture a considerably greater degree of data variation caused both by larger technical and pathological variance in the datasets.

Fourth, vessel segmentation provides a unique challenge owing to the complex nature of the vascular structure. Arterial brain vessels comprise around 1.5% of the input volume and therefore lead to an extreme class imbalance in the dataset. To mitigate this effect, a DSC loss function was used to neglect true negatives and subsampling of the training patches was applied so that 50% of all patches are centered around vessels ensuring representative and fair sampling. However, to properly evaluate vessel segmentation models, the performance measures must also reflect the vessel properties. Given ground-truth segmentations, accuracy, precision, and recall are the most frequent measures but numerous other measures exist (10,28). DSC is the most widely used metric and is therefore essential for examining the results with respect to the literature. Nonetheless, distance measures such as AVD or 95HD have a distinct advantage over DSC in the context of vessels. Both DSC and HD measures are based on voxel-to-voxel comparisons between the segmented vessels and the ground-truth labels, without considering the continuity properties of the brain vasculature. However, since HD penalizes with respect to distance from the labeling it results in an implicit preference for (approximated) continuous structures. Previous studies have suggested HD measures as more useful to assess the performance of vessel segmentations (Taha and Hanbury, 2015). Thus, we assessed model performance based on five different measures: Precision, recall, DSC, AVD and 95HD. This selection of metrics gives a comprehensive and complete overview of model performance as well as enables the presented state-of-the-art to be benchmarked thoroughly - i.e. beyond DSC - against future research.

Fifth, one of the major strengths of our study is the validation of the models in patients with pathological conditions. Many pathologies are usually represented only by a small number of voxels compared to the full number of vessel voxels per patient. Some pathologies are present only in a small number of patients and are rare even in large samples. Therefore, these anomalies cannot be reflected properly by numerical performance measures but must be assessed visually. Thus, we performed a thorough visual analysis of the results by a stroke imaging expert. Our findings indicated that the BRAVE-NET architecture outperformed the other models and was less likely to misclassify anomalies such as proteinaceous fluid or laminar cortical necrosis. Nevertheless, the ratio of misclassifications, albeit often of minor markedness, was still 40% of all images. This underlines the importance of qualitative visual assessments to examine segmentation models for clinical applications. When quantitative metrics begin to plateau and stop showing meaningful differences, visual assessment is necessary to make clinically relevant distinctions. We, therefore, advise future studies to validate all proposed models both quantitatively and qualitatively, especially for the development of clinical applications.

To summarize, the presented framework add to the literature through the unique combination of high performance, full automation and robust validation. It thus has the largest applicability for the clinical setting so far.

The excellent performance of BRAVE-NET can also be attributed to architecture design choices. CNNs and encoder-decoder Unet variations, in particular, were proven in recent years as the state-of-the-art method for semantic segmentation including medical imaging applications (Milletari et al., 2016; Badrinarayanan et al., 2017; Dong et al., 2017; Alom et al., 2018; Norman et al., 2018). This success was further demonstrated in vessel segmentation and specifically in brain vessel segmentation problems (Chen et al., 2017; Alom et al., 2018; Huang et al., 2018; Livne et al., 2019a; Yue et al., 2019). The utilization of the Unet as a baseline architecture was, therefore, a natural choice. Nonetheless, the classic Unet architecture was previously shown to be prone to errors in the presence of visible pathologies especially in segmenting small vessels and with regards to the skull (Livne et al., 2019a). To target these issues we aimed to improve spatial information integration using multiscale- and 3-D variants. Multiscale architectures - i.e. fusion of information from multiple resolutions - were shown to be beneficial in boundary detection and segmentation problems (Choi et al.; Kokkinos, 2015; Kamnitsas et al., 2017; Stawiaski, 2017). We implemented multiscaling following a common practice across applications, namely employing an additional encoder path of the architecture operating on lower resolution. This so - called context path, allowed to capture the broader anatomical context of the vessels and particularly target the detection of small vessels. Multiscale integration was also important to tackle the challenge of unmasked images: it allowed distinguishing larger anatomical structures such as the skull and neck areas from the regions with brain and brain-supplying vessels while maintaining high resolution of the segmented vessels. The down-sampling within the context path enabled to avoid redundant extraction of features while maintaining corresponding spatial integrity of the parallel paths. Multiscale models yielded improved performance in all measures compared to the baseline Unet, corroborating the advantage of multiscaling in semantic segmentation tasks. Moreover, deep supervision in CNNs was previously shown to be advantageous as a strong “regularization” for classification accuracy and learned features and tackling problematic convergence behavior (Lee et al., 2015). Different variations of deep CNNs were described in the literature and demonstrated improved results and faster convergence (Lee et al., 2015; Szegedy et al., 2015; Wang et al., 2015a; Dou et al., 2016). In our study, deep supervision improved performance only in combination with the multiscaling approach.

Our study has several recognized limitations. First, while the dataset in this study was acquired from multiple scanners and patients exhibited a large spectrum of steno-occlusive cerebrovascular disease, the data heterogeneity was still limited. It is highly warranted to specifically include in future work datasets with other pathologies such as aneurysms, other MR scanner- and MR imaging sequence types and other imaging modalities such as CT. Here, transfer learning approaches seem beneficial. Second, it was beyond the scope of this work to integrate all potentially beneficial architecture modifications. Interesting further future modifications could be skip connections between convolutional blocks to explicitly exploit multiscale features (Zhou et al., 2020) or densely connected convolutional blocks to enhance feature propagation through encoding levels (Zhang et al., 2018). Third, due to well recognized resource constraints only one fold was reviewed visually. It is a general challenge for this kind of research that quality characteristics such as cross validation and large data sets lead to an increasing amount of images that require highly time-consuming visual review. It is unlikely that reviewing more folds would lead to major changes in the visual assessment, but it cannot be ruled out with certainty.

## Conclusion

We present a fully automated deep learning framework for TOF-MRA brain vessel segmentation. We utilize multiscaling for better spatial information integration and deep supervision for improving model convergence. We employ a multi-scanner dataset of yet unmatched size and range of pathologies in the published literature. We also ensure a high degree of robustness of our framework through extensive quantitative and qualitative evaluation. We explicitly analyze and communicate model performance in presence of relevant pathologies to further increase applicability in the clinical setting. Our results thus constitute a new state-of-the-art for fully automated frameworks for brain vessel segmentation. Our work strongly facilitates and promotes the usability of automated vessel segmentation in the clinical setting and can be swiftly translated to the development of cerebrovascular biomarkers for clinical applications.

## Data Availability

Due to data protection laws, the imaging data used in this study cannot be published at the current time point.

## References

Abadi, M., Agarwal, A., Barham, P., Brevdo, E., Chen, Z., Citro, C., et al. TensorFlow: Large-Scale Machine Learning on Heterogeneous Distributed Systems. 19.

Alom, M. Z., Hasan, M., Yakopcic, C., Taha, T. M., and Asari, V. K. (2018). Recurrent Residual Convolutional Neural Network based on U-Net (R2U-Net) for Medical Image Segmentation. ArXiv180206955 Cs. Available at: http://arxiv.org/abs/1802.06955 [Accessed October 3, 2019].

Babin, D., Pižurica, A., Vylder, J. D., Vansteenkiste, E., and Philips, W. (2013). Brain blood vessel segmentation using line-shaped profiles. Phys. Med. Biol. 58, 8041–8061. doi:10.1088/0031-9155/58/22/8041.

Badrinarayanan, V., Kendall, A., and Cipolla, R. (2017). SegNet: A Deep Convolutional Encoder-Decoder Architecture for Image Segmentation. IEEE Trans. Pattern Anal. Mach. Intell. 39, 2481–2495. doi:10.1109/TPAMI.2016.2644615.

Chen, L., Xie, Y., Sun, J., Balu, N., Mossa-Basha, M., Pimentel, K., et al. (2017). Y-net: 3D intracranial artery segmentation using a convolutional autoencoder. ArXiv171207194 Cs Eess. Available at: http://arxiv.org/abs/1712.07194 [Accessed September 19, 2019].

Chen, L.-C., Papandreou, G., Kokkinos, I., Murphy, K., and Yuille, A. L. (2014). Semantic Image Segmentation with Deep Convolutional Nets and Fully Connected CRFs. ArXiv14127062 Cs. Available at: http://arxiv.org/abs/1412.7062 [Accessed October 18, 2019].

Choi, Y., Kwon, Y., Lee, H., Paik, M. C., and Won, J.-H. 3D Multiscale Residual U-Net Architecture for Brain Lesion Segmentation. 27.

Dengler, N. F., Madai, V. I., Wuerfel, J., von Samson-Himmelstjerna, F. C., Dusek, P., Niendorf, T., et al. (2016). Moyamoya Vessel Pathology Imaged by Ultra-High-Field Magnetic Resonance Imaging at 7.0?T. J. Stroke Cerebrovasc. Dis. Off. J. Natl. Stroke Assoc. 25, 1544–1551. doi:10.1016/j.jstrokecerebrovasdis.2016.01.041.

Dong, H., Yang, G., Liu, F., Mo, Y., and Guo, Y. (2017). Automatic Brain Tumor Detection and Segmentation Using U-Net Based Fully Convolutional Networks. in Medical Image Understanding and Analysis Communications in Computer and Information Science., eds. M. Valdés Hernández and V. González-Castro (Springer International Publishing), 506–517.

Dou, Q., Chen, H., Jin, Y., Yu, L., Qin, J., and Heng, P.-A. (2016). 3D Deeply Supervised Network for Automatic Liver Segmentation from CT Volumes. in Medical Image Computing and Computer-Assisted Intervention – MICCAI 2016 Lecture Notes in Computer Science., eds. S. Ourselin, L. Joskowicz, M. R. Sabuncu, G. Unal, and W. Wells (Springer International Publishing), 149–157.

Dutra, B. G., Tolhuisen, M. L., Alves, H. C. B. R., Treurniet, K. M., Kappelhof, M., Yoo, A. J., et al. (2019). Thrombus Imaging Characteristics and Outcomes in Acute Ischemic Stroke Patients Undergoing Endovascular Treatment. Stroke 50, 2057–2064. doi:10.1161/STROKEAHA.118.024247.

Folle, L., Vesal, S., Ravikumar, N., and Maier, A. (2019). Dilated Deeply Supervised Networks for Hippocampus Segmentation in MRI. in Bildverarbeitung für die Medizin 2019 Informatik aktuell., eds. H. Handels, T. M. Deserno, A. Maier, K. H. Maier-Hein, C. Palm, and T. Tolxdorff (Springer Fachmedien Wiesbaden), 68–73.

Glorot, X., and Bengio, Y. (2010). Understanding the difficulty of training deep feedforward neural networks. in Proceedings of the Thirteenth International Conference on Artificial Intelligence and Statistics, 249–256. Available at: http://proceedings.mlr.press/v9/glorot10a.html [Accessed October 18, 2019].

Gutierrez Jose, Cheung Ken, Bagci Ahmet, Rundek Tatjana, Alperin Noam, Sacco Ralph L., et al. (2015). Brain Arterial Diameters as a Risk Factor for Vascular Events. J. Am. Heart Assoc. 4, e002289. doi:10.1161/JAHA.115.002289.

Ho, C. W. L., Soon, D., Caals, K., and Kapur, J. (2019). Governance of automated image analysis and artificial intelligence analytics in healthcare. Clin. Radiol. 74, 329–337. doi:10.1016/j.crad.2019.02.005.

Hofmanninger, J., Prayer, F., Pan, J., Rohrich, S., Prosch, H., and Langs, G. (2020). Automatic lung segmentation in routine imaging is a data diversity problem, not a methodology problem. ArXiv200111767 Phys. Stat. Available at: http://arxiv.org/abs/2001.11767 [Accessed March 24, 2020].

Hotter, B., Pittl, S., Ebinger, M., Oepen, G., Jegzentis, K., Kudo, K., et al. (2009). Prospective study on the mismatch concept in acute stroke patients within the first 24 h after symptom onset - 1000Plus study. BMC Neurol. 9, 60. doi:10.1186/1471-2377-9-60.

Huang, Q., Sun, J., Ding, H., Wang, X., and Wang, G. (2018). Robust liver vessel extraction using 3D U-Net with variant dice loss function. Comput. Biol. Med. 101, 153–162. doi:10.1016/j.compbiomed.2018.08.018.

Ioffe, S., and Szegedy, C. (2015). Batch Normalization: Accelerating Deep Network Training by Reducing Internal Covariate Shift. ArXiv150203167 Cs. Available at: http://arxiv.org/abs/1502.03167 [Accessed October 18, 2019].

Kamnitsas, K., Ledig, C., Newcombe, V. F. J., Simpson, J. P., Kane, A. D., Menon, D. K., et al. (2017). Efficient multi-scale 3D CNN with fully connected CRF for accurate brain lesion segmentation. Med. Image Anal. 36, 61–78. doi:10.1016/j.media.2016.10.004.

Kingma, D. P., and Ba, J. (2014). Adam: A Method for Stochastic Optimization. ArXiv14126980 Cs. Available at: http://arxiv.org/abs/1412.6980 [Accessed October 18, 2019].

Kokkinos, I. (2015). Pushing the Boundaries of Boundary Detection using Deep Learning. ArXiv151107386 Cs. Available at: http://arxiv.org/abs/1511.07386 [Accessed October 15, 2019].

Lee, C., Xie, S., Gallagher, P., Zhang, Z., and Tu, Z. (2015). Deeply-Supvervised Nets. in, x562–570.

Lesage, D., Angelini, E. D., Bloch, I., and Funka-Lea, G. (2009). A review of 3D vessel lumen segmentation techniques: Models, features and extraction schemes. Med. Image Anal. 13, 819–845. doi:10.1016/j.media.2009.07.011.

Livne, M., Rieger, J., Aydin, O. U., Taha, A. A., Akay, E. M., Kossen, T., et al. (2019a). A U-Net Deep Learning Framework for High Performance Vessel Segmentation in Patients With Cerebrovascular Disease. Front. Neurosci. 13. doi:10.3389/fnins.2019.00097.

Livne, M., Rieger, J., Aydin, O. U., Taha, A. A., Akay, E. M., Kossen, T., et al. (2019b). Model Data for “A U-Net Deep Learning Framework for High Performance Vessel Segmentation in Patients With Cerebrovascular Disease.” doi:10.5281/zenodo.3433818.

Long, J., Shelhamer, E., and Darrell, T. (2014). Fully Convolutional Networks for Semantic Segmentation. ArXiv14114038 Cs. Available at: http://arxiv.org/abs/1411.4038 [Accessed October 18, 2019].

Martin, S. Z., Madai, V. I., von Samson-Himmelstjerna, F. C., Mutke, M. A., Bauer, M., Herzig, C. X., et al. (2015). 3D GRASE Pulsed Arterial Spin Labeling at Multiple Inflow Times in Patients with Long Arterial Transit Times: Comparison with Dynamic Susceptibility-Weighted Contrast-Enhanced MRI at 3 Tesla. J. Cereb. Blood Flow Metab. 35, 392–401. doi:10.1038/jcbfm.2014.200.

Meijs, M., Patel, A., Leemput, S. C. van de, Prokop M., Dijk, E. J. van, Leeuw, F.-E. de, et al. (2017). Robust Segmentation of the Full Cerebral Vasculature in 4D CT of Suspected Stroke Patients. Sci. Rep. 7, 1–12. doi:10.1038/s41598-017-15617-w.

Milletari, F., Navab, N., and Ahmadi, S.-A. (2016). V-Net: Fully Convolutional Neural Networks for Volumetric Medical Image Segmentation. ArXiv160604797 Cs. Available at: http://arxiv.org/abs/1606.04797 [Accessed October 3, 2019].

Mitchell, M., Wu, S., Zaldivar, A., Barnes, P., Vasserman, L., Hutchinson, B., et al. (2019). Model Cards for Model Reporting. in Proceedings of the Conference on Fairness, Accountability, and Transparency FAT* ‘19. (Atlanta, GA, USA: Association for Computing Machinery), 220–229. doi:10.1145/3287560.3287596.

Moccia, S., De Momi, E., El Hadji, S., and Mattos, L. S. (2018). Blood vessel segmentation algorithms — Review of methods, datasets and evaluation metrics. Comput. Methods Programs Biomed. 158, 71–91. doi:10.1016/j.cmpb.2018.02.001.

Murray, N. M., Unberath, M., Hager, G. D., and Hui, F. K. (2019). Artificial intelligence to diagnose ischemic stroke and identify large vessel occlusions: a systematic review. J. NeuroInterventional Surg., neurintsurg-2019-015135. doi:10.1136/neurintsurg-2019-015135.

Ni, J., Wu, J., Wang, H., Tong, J., Chen, Z., Wong, K. K. L., et al. (2020). Global channel attention networks for intracranial vessel segmentation. Comput. Biol. Med. 118, 103639. doi:10.1016/j.compbiomed.2020.103639.

Norman, B., Pedoia, V., and Majumdar, S. (2018). Use of 2D U-Net Convolutional Neural Networks for Automated Cartilage and Meniscus Segmentation of Knee MR Imaging Data to Determine Relaxometry and Morphometry. Radiology 288, 177–185. doi:10.1148/radiol.2018172322.

Passat, N., Ronse, C., Baruthio, J., Armspach, J.-P., Maillot, C., and Jahn, C. (2005). Region-growing segmentation of brain vessels: An atlas-based automatic approach. J. Magn. Reson. Imaging 21, 715–725. doi:10.1002/jmri.20307.

Patel, T. R., Paliwal, N., Jaiswal, P., Waqas, M., Mokin, M. M.d A. H. S., et al. (2020). Multi-resolution CNN for brain vessel segmentation from cerebrovascular images of intracranial aneurysm: a comparison of U-Net and DeepMedic. in Medical Imaging 2020: Computer-Aided Diagnosis (International Society for Optics and Photonics), 113142W. doi:10.1117/12.2549761.

Ronneberger, O., Fischer, P., and Brox, T. (2015). U-Net: Convolutional Networks for Biomedical Image Segmentation. in Medical Image Computing and Computer-Assisted Intervention – MICCAI 2015 Lecture Notes in Computer Science., eds. N. Navab, J. Hornegger, W. M. Wells, and A. F. Frangi (Springer International Publishing), 234–241.

Santos, E. M. M., Marquering, H. A., den Blanken, M. D., Berkhemer, O. A., Boers, A. M. M., Yoo, A. J., et al. (2016). Thrombus Permeability Is Associated With Improved Functional Outcome and Recanalization in Patients With Ischemic Stroke. Stroke 47, 732–741. doi:10.1161/STROKEAHA.115.011187.

Srivastava, N., Hinton, G., Krizhevsky, A., Sutskever, I., and Salakhutdinov, R. (2014). Dropout: A Simple Way to Prevent Neural Networks from Overfitting. J. Mach. Learn. Res. 15, 1929–1958.

Stawiaski, J. (2017). A Multiscale Patch Based Convolutional Network for Brain Tumor Segmentation. ArXiv abs/1710.02316.

Szegedy, C., Liu, W., Jia, Y., Sermanet, P., Reed, S., Anguelov, D., et al. (2015). Going Deeper With Convolutions. in, 1–9. Available at: https://www.cv-foundation.org/openaccess/content_cvpr_2015/html/Szegedy_Going_Deeper_With_2015_CVPR_paper.html [Accessed October 5, 2019].

Taha, A. A., and Hanbury, A. (2015). Metrics for evaluating 3D medical image segmentation: analysis, selection, and tool. BMC Med. Imaging 15. doi:10.1186/s12880-015-0068-x.

Taher, F., Soliman, A., Kandil, H., Mahmoud, A., Shalaby, A., Gimelrfarb, G., et al. (2020). Accurate Segmentation of Cerebrovasculature from TOF-MRA Images using Appearance Descriptors. IEEE Access, 1–1. doi:10.1109/ACCESS.2020.2982869.

Tetteh, G., Efremov, V., Forkert, N. D., Schneider, M., Kirschke, J., Weber, B., et al. (2018). DeepVesselNet: Vessel Segmentation, Centerline Prediction, and Bifurcation Detection in 3-D Angiographic Volumes. ArXiv180309340 Cs. Available at: http://arxiv.org/abs/1803.09340 [Accessed July 14, 2018].

Turc, G., Bhogal, P., Fischer, U., Khatri, P., Lobotesis, K., Mazighi, M., et al. (2019). European Stroke Organisation (ESO) – European Society for Minimally Invasive Neurological Therapy (ESMINT) Guidelines on Mechanical Thrombectomy in Acute Ischaemic StrokeEndorsed by Stroke Alliance for Europe (SAFE). Eur. Stroke J. 4, 6–12. doi:10.1177/2396987319832140.

Ultrahigh-Field MRI in Human Ischemic Stroke – a 7 Tesla Study Available at: https://journals.plos.org/plosone/article?id=10.1371/journal.pone.0037631 [Accessed January 14, 2020].

Wang, L., Lee, C.-Y., Tu, Z., and Lazebnik, S. (2015a). Training Deeper Convolutional Networks with Deep Supervision. ArXiv150502496 Cs. Available at: http://arxiv.org/abs/1505.02496 [Accessed October 5, 2019].

Wang, R., Li, C., Wang, J., Wei, X., Li, Y., Zhu, Y., et al. (2015b). Threshold segmentation algorithm for automatic extraction of cerebral vessels from brain magnetic resonance angiography images. J. Neurosci. Methods 241, 30–36. doi:10.1016/j.jneumeth.2014.12.003.

WHO EMRO | Stroke, Cerebrovascular accident | Health topics Available at: http://www.emro.who.int/health-topics/stroke-cerebrovascular-accident/index.html [Accessed September 20, 2019].

Yoo, J., Baek, J.-H., Park, H., Song, D., Kim, K., Hwang, I. G., et al. (2018). Thrombus Volume as a Predictor of Nonrecanalization After Intravenous Thrombolysis in Acute Stroke. Stroke 49, 2108–2115. doi:10.1161/STROKEAHA.118.021864.

Yue, K., Zou, B., Chen, Z., and Liu, Q. (2019). Retinal vessel segmentation using dense U-net with multiscale inputs. J. Med. Imaging 6, 034004. doi:10.1117/1.JMI.6.3.034004.

Yushkevich, P. A., Piven, J., Hazlett, H. C., Smith, R. G., Ho, S., Gee, J. C., et al. (2006). User-guided 3D active contour segmentation of anatomical structures: Significantly improved efficiency and reliability. NeuroImage 31, 1116–1128. doi:10.1016/j.neuroimage.2006.01.015.

Zhang, J., Jin, Y., Xu, J., Xu, X., and Zhang, Y. (2018). MDU-Net: Multi-scale Densely Connected U-Net for biomedical image segmentation. ArXiv181200352 Cs. Available at: http://arxiv.org/abs/1812.00352 [Accessed May 23, 2019].

Zhou, Z., Siddiquee, M. M. R., Tajbakhsh, N., and Liang, J. (2020). UNet++: Redesigning Skip Connections to Exploit Multiscale Features in Image Segmentation. ArXiv191205074 Cs Eess. Available at: http://arxiv.org/abs/1912.05074 [Accessed March 24, 2020].

